# Deep Learning-Based Pretreatment Cardiovascular Risk Stratification in Women with Breast Cancer

**DOI:** 10.64898/2026.07.26.26350307

**Authors:** Mehdi Dehghan Manshadi, Narges Manouchehri, Laila Hubbert, Annelie Liljegren, Ali Manouchehrinia, Christina Linder-Stragliotto, Johanna Rantala, Elham Hedayati, Narsis A. Kiani

## Abstract

**Background:** Cardiovascular disease is a leading non-cancer cause of morbidity and mortality among breast cancer (BC) survivors. Existing cardiovascular risk tools are not tailored to cancer populations and often rely on cardiology investigations or treatment details unavailable at the initial oncology visit, limiting their use for early referral decisions.

**Methods:** We conducted a registry-based cohort study including 17,051 women diagnosed with BC in stage I-III or ductal carcinoma in situ in the Stockholm-Gotland region (2008-2019). Using only pre-treatment information routinely available to oncologists, such as demographics, cancer characteristics, planned cancer treatment, baseline comorbidities, medications, and healthcare utilization, we trained and validated a deep learning-based competing-risk model to predict 1-year major adverse cardiovascular events (MACE) risk, accounting for non-cardiovascular death as a competing outcome. Model performance was evaluated using a 3-fold CV and time-dependent concordance indices. Fine-Gray subdistribution hazard models were used to aid interpretability.

**Results:** The model demonstrated strong and stable discrimination across validation folds, with a median c-index of 0.84 for 1-year MACE prediction and 0.90 for the competing risk. Key contributors to predictive performance included age at BC diagnosis, prior cardiovascular disease, healthcare utilization patterns, cancer stage, and specific medication profiles. Several predictors with modest marginal hazard ratios in the Fine-Gray model contributed substantially through nonlinear effects and interactions.

**Conclusions:** Our model with a deep learning-based competing-risk structure and using only pre-treatment, oncology-accessible data enables accurate short-term cardiovascular risk stratification in women with BC and may support targeted cardio-oncology referral prior to initiation of systemic therapy.

## Introduction

Cardiovascular disease (CVD) has emerged as a major determinant of long-term outcomes in patients with cancer. It is now a leading non-cancer cause of morbidity and mortality in breast cancer (BC) survivors. Large population-based and cohort studies consistently show that patients with BC experience an excess burden of heart failure, ischemic events, and stroke compared with individuals without cancer, driven both by shared risk factors and by cardiotoxic cancer therapies^1–3^. This growing population of BC survivors has created an urgent need for cardio-oncology tools that can identify patients at high risk for CVD early in the treatment pathway, ideally before systemic therapy is initiated.

Several widely used general-population risk scores, such as QRISK3, SCORE2, PCP-HF, and CHARGE-AF, provide robust estimation of atherosclerotic CVD, heart failure, or atrial fibrillation risk in community cohorts^4–7^. However, these calculators were not derived in cancer populations, typically assume stable long-term risk factor profiles, and often require standardized measurements (e.g., lipid panels, sometimes ECG variables) that may not be systematically available at the time of an oncology visit. Moreover, they do not account for cancer-specific characteristics, upcoming oncologic therapies, or treatment-related cardiotoxicity, and they generally treat CVD as a single composite endpoint without considering the competing risk of cancer-related mortality.

To address these limitations, multiple cardio-oncology-specific scores have been developed for BC patients receiving potentially cardiotoxic therapies. Early models by Ezaz et al. and Fogarassy et al. used claims or registry data to predict heart failure or cardiomyopathy after anthracycline and trastuzumab exposure, relying on traditional regression-based risk scores built from age, comorbidities, and chemotherapy categories ^8,9^. Subsequent models have leveraged imaging data and more granular treatment information, including scores based on baseline and on-treatment Left Ventricular Ejection Fraction (LVEF) to identify low-risk trastuzumab-treated patients ^10^, regression-based risk scores that integrate demographics, conventional cardiovascular risk factors (CVRFs), anthracycline dose, radiotherapy laterality, and combined modality therapy to estimate 3-10-year major adverse cardiovascular events (MACE) risk ^11^, and echocardiography-based models that combine global longitudinal strain, left ventricular (LV) dimensions, heart rate, and planned anthracycline/HER2-targeted therapy to predict cancer therapy-related cardiac dysfunction prior to chemotherapy initiation^12^. Despite these advances, such tools typically rely on specialized cardiology investigations (echocardiography, strain imaging, and sometimes biomarkers), detailed information on delivered oncologic regimens, and longitudinal on-treatment measurements. Therefore, these tools do not directly address the earlier decision point at which an oncologist must determine whether to involve a cardiologist before ordering or starting systemic therapy.

Large-scale electronic health record (EHR) datasets and machine-learning methods have recently been leveraged to develop cancer-specific cardiovascular risk models. Multi-modal frameworks that integrate EHR data with patient-reported lifestyle factors and social determinants of health have been used to predict multiple CVD outcomes over a mid-to long-range horizon (i.e., 10-year) in BC cohorts, typically combining univariate Cox-based feature screening with regularized Cox models and random forests evaluated by concordance index (c-index) and time-dependent area under the curve (AUC)^12^. Other work has derived cancer-specific mid-to long-range CVD risk equations for breast, colorectal, and lung cancer using EHR and registry data, employing XGBoost-guided feature selection followed by parsimonious logistic regression scores that outperform general-population tools such as PCE^13^, PREVENT^14^, and SCORE2^15^. However, these approaches generally require detailed treatment and laboratory data, formulate risk as a binary “event within 10 years” outcome, and focus on long-term post-diagnosis risk rather than the pre-treatment referral decisions faced in routine cardio-oncology care.

Deep-learning approaches have further expanded the methodological landscape. Longitudinal EHR-based sequence models with trainable decay terms that incorporate NLP-derived tumor features have achieved AUC of approximately 0.72-0.95 for predicting multiple cardiac endpoints in women with breast cancer when 12-24 months of history are available^16^. Other work applying classical and deep machine-learning classifiers to real-world breast-cancer cohorts has reported AUC in the 0.78-0.90 range for early post-treatment cardiovascular events following chemotherapy, targeted therapy, endocrine therapy, or radiotherapy ^17^. In the immune-checkpoint-inhibitor setting, multimodal fusion AI models that integrate clinical, laboratory, ECG, echocardiographic, and treatment data have shown good discrimination, with AUC values in the low 0.7 to low 0.8 range. These models consistently outperform single-modality approaches^18^. In addition, XGBoost-based models trained on large registries such as CancerLinQ, together with regression-based models developed using TriNetX data, have demonstrated moderate but clinically useful performance for predicting immune-checkpoint-inhibitor-related cardiac events^19,20^. Collectively, these studies highlight the promise of multimodal EHR- and imaging-based AI for cardio-oncology, but they typically rely on rich longitudinal data, detailed treatment patterns, and cardiology-specific tests that are often not available at the initial oncology consultation.

Despite these advances, a critical gap remains because existing cardio-oncology tools are not designed to support the baseline decision of whether a newly diagnosed BC patient needs to be referred to a cardiologist before cancer-directed therapy, using only information available in the oncology clinic. Given rising patient volumes and limited cardio-oncology capacity, it is neither feasible nor desirable to refer all patients, underscoring the need for targeted, risk-based referral strategies. In this registry-based matched cohort study within the Stockholm-Gotland region in Sweden, we address this gap by training and validating a deep learning-based prognostic model that estimates the risk of 1-year MACE ts in women with BC using only pre-treatment information typically accessible to oncologists, namely demographics, cancer characteristics, and baseline comorbidities and medications, while deliberately excluding echocardiography, ECG-derived metrics, specialized cardiac biomarkers, and chemotherapy or radiotherapy doses. The model provides individualized, time-dependent risk estimates for MACE, accounting for non-cardiovascular death as a competing risk. This model is intended as a practical decision-support tool to help identify patients who may benefit from cardiology referral prior to initiation of systemic cancer therapy.

## Methods

The study was designed as a retrospective register-based cohort analysis. The cohort included 17,051 women diagnosed with BC (stages I-III and ductal carcinoma in situ [DCIS]) in the Stockholm-Gotland region of Sweden between January 1, 2008, and December 31, 2019.

Patients with unknown TNM, cancer stage, or known BRCA1/2 germline mutation were excluded from the study (Fig 1).

Data from regional and national quality registries provided detailed information on pre-existing CVD, CVRFs, pre-index diagnostic history, planned cancer treatments, demographics, prescribed medications, and underlying and contributing causes of death. The data sources and linkage procedures have been described previously^21^. Cardiovascular outcomes were identified from both inpatient and outpatient records using International Classification of Diseases (ICD) codes and registry-defined interventions.

**Fig1**.

This study was approved by the Swedish Ethical Review Board (2018/2669-31/2, amendment 2019-00540, 2020-07086 and 2024-00960-02) and adhered to the STROBE guidelines^22^ and the Declaration of Helsinki^23^. According to Swedish legislation, written informed consent is not required from individuals included in national quality registries for the use of their data in healthcare research.

### Outcomes

The primary outcome was a composite MACE endpoint within the first year after BC diagnosis. Here, the composite MACE (hereafter referred to as MACE) was defined as non-fatal myocardial infarction (I21), non-fatal heart failure (I50), non-fatal stroke (I61), or cardiovascular death. Cardiovascular death was defined as any death in which an ICD-10 code from I00-I99 was recorded as the underlying or a contributing cause of death in the national Cause of Death Register. The competing outcome was death from non-cardiovascular causes within the first year after BC diagnosis.

### Covariates and features

Demographics, tumor characteristics, planned cancer treatment, pre-index diagnostic history, prescribed medications, and healthcare utilization features were obtained from regional and national quality registries, restricted to information recorded up to the index BC diagnosis.

Planned cancer treatment was defined as the first intended oncologic management strategy recorded after diagnosis (treatment intent and modality), reflecting the care plan prior to completion of therapy and before later modifications due to toxicity, progression, or patient preference. These planned treatment modalities included all planned therapies for chemotherapy, radiotherapy, anti-HER2-targeted therapy, and endocrine (hormonal) therapy. Pre-index diagnostic histories were retrieved using ICD-10 codes and grouped according to the main ICD-10 chapter categories (e.g., diseases of the circulatory system, endocrine and metabolic diseases, respiratory diseases) to construct comorbidity and disease-history variables. In addition to the broad group of diseases of the circulatory system (ICD-10 codes I00-I99), we defined a subgroup of specific ICD-10 codes (Supplementary Table S1) corresponding to selected cardiovascular diseases, referred to as *Pre-existing CVD*. Prescribed medications were mapped to Anatomical Therapeutic Chemical (ATC) codes truncated at level 2. Additional healthcare utilization features were derived, including the number of outpatient visits, the maximum duration of hospitalization, and the age at the first recorded circulatory system disease. For clarity, features were grouped into five major categories: Basics, Planned Treatment, ATCs, RFs, ICDs, and Extracted. A detailed description of the features included in each category, along with their abbreviations, is provided in Supplementary Table S2.

### Risk prediction model

For predicting the 1-year MACE after BC diagnosis, we borrowed the main idea from the DeepHit model^24^, a deep learning-based approach that explicitly accounts for competing risks in time-to-event analyses. DeepHit enables flexible modeling of nonlinear effects and higher-order interactions among CVRFs, tumor characteristics, and treatments within a competing-risks framework.

### Model architecture

The neural network architecture consisted of a shared multilayer perceptron (MLP) with hidden layers of sizes 30, 15, and 5, followed by cause-specific subnetworks with hidden layers of sizes 5 and 10 for each event type. Rectified Linear Unit (ReLU) activations were used for all hidden layers. The output layer employed a softmax activation over discretized time intervals, producing normalized, time-dependent probability mass functions for all event types (Fig 2).

**Fig 2**

Although the covariate space was moderate, the inputs were sparse binary indicators, and the outcome was comparatively sparse (∼2.5%), while the model must learn a structured joint distribution over discrete time and competing events. We therefore used an encoder-decoder architecture to (i) learn a compact latent representation that aggregates weak and infrequent signals and captures non-linear interactions among binary covariates, and (ii) decode this shared representation into event-specific time distributions with parameter sharing across time bins and event types. Compared with a single feed-forward network that directly maps covariates to all time-event outputs, this factorized design imposes an implicit regularization on the predicted time-event surface, which can improve training stability and data efficiency in the presence of limited event information and censoring.

### Model training and loss function

The model was trained using the combined likelihood and ranking loss functions proposed in the original DeepHit framework. The likelihood component (eq. 1) encourages the model to assign a high probability to the observed event outcome by maximizing the probability mass at the observed event time and event type for uncensored individuals. For censored individuals, it increases the predicted survival probability beyond the censoring time by allocating probability mass to later times.

**Eq. 1.**

The other component of the loss function is a ranking loss (Eq. 2), which targets discrimination by enforcing the correct risk ordering among comparable subjects. For a given event type, individuals who experience the event earlier are encouraged to have a higher predicted cumulative incidence at that time than individuals who are still event-free and at risk. Together, the likelihood and ranking terms promote both calibration to censored time-to-event data and strong discriminative performance in competing-risk prediction.

**Eq. 2.**

Model evaluation was performed using 3-fold cross-validation (CV). The full cohort was randomly partitioned into three mutually exclusive folds, with two folds used for model training and the remaining fold used for validation in each iteration. Predictive performance was then evaluated by assessing how accurately the model predicted the occurrence of MACE within the first year after BC diagnosis in the validation cohorts. Discrimination was quantified using the time-dependent c-index^25^.

### Feature importance

To facilitate interpretation and compare the relative contribution of individual predictors across modeling frameworks, we computed model-agnostic feature importance using permutation; for each covariate, its values were repeatedly permuted in the test data 100 times, and the resulting change in predictive performance was quantified. A larger average performance drop is interpreted as indicating greater importance for 1-year MACE prediction.

### Effect of covariates on the risk

While our model was used for risk prediction due to its ability to capture nonlinear effects and complex interactions in the presence of competing risks, we additionally fitted the Fine-Gray subdistribution hazards models^26^ to obtain direction and magnitude of associations between covariates and 1-year MACE. The Fine-Gray models provide interpretable subdistribution hazard ratios under proportional hazards assumptions, thereby facilitating clinical interpretation of covariate effects.

### Statistical analysis

Pairwise correlations between features were computed using Pearson’s correlation coefficient. Deep learning-based model fitting were performed using Python (version 3.12)^27^. Data preprocessing, analyses related to Fine-Gray model, and visualizations were conducted in R version 4.5.1^28^ and RStudio 5.1^29^.

## Results

### The prevalence of the main outcome is less than one percent

The baseline characteristics and demographic features of the study cohort are summarized in Table 1. The primary outcome occurred in 447 patients, representing a relatively rare event for model development and prediction. The incidence of events during the first year shows a marked increase in the primary outcome, underscoring the importance of focusing on 1-year MACE risk stratification as the primary objective of this study.

**Table 1.**

### The model shows acceptable prediction power for both risks

The discriminative performance of the model was evaluated using a 3-fold CV (Table 2). Across the three folds, the model demonstrated consistently high discrimination for the main risk, with a median c-index of 84.0% across folds. Similarly, discrimination for the competing risk was high, with a median c-index of 89.8% across folds. These results indicate stable and robust discriminative ability of our model for individual risk stratification in the presence of competing events.

**Table 2.** The c-index for each of the 3-fold CVs.

|  | 1st CV | 2nd CV | 3rd CV |
| --- | --- | --- | --- |
| Main risk | 83.2 | 84.4 | 84.0 |
| Competing risk | 90.1 | 84.3 | 89.8 |

### Features included in the model showed no significant correlation

Among all features, only ICDs were excluded from the final model, as their inclusion did not improve predictive performance. Pairwise correlation analysis showed no strong associations between features, indicating that the observed predictive performance was unlikely to be driven by redundant information (Figure 3). The strongest observed correlation was between Diabetes and ATC A10, medications related to diabetes, with a correlation coefficient < 0.7.

**Figure 3.**
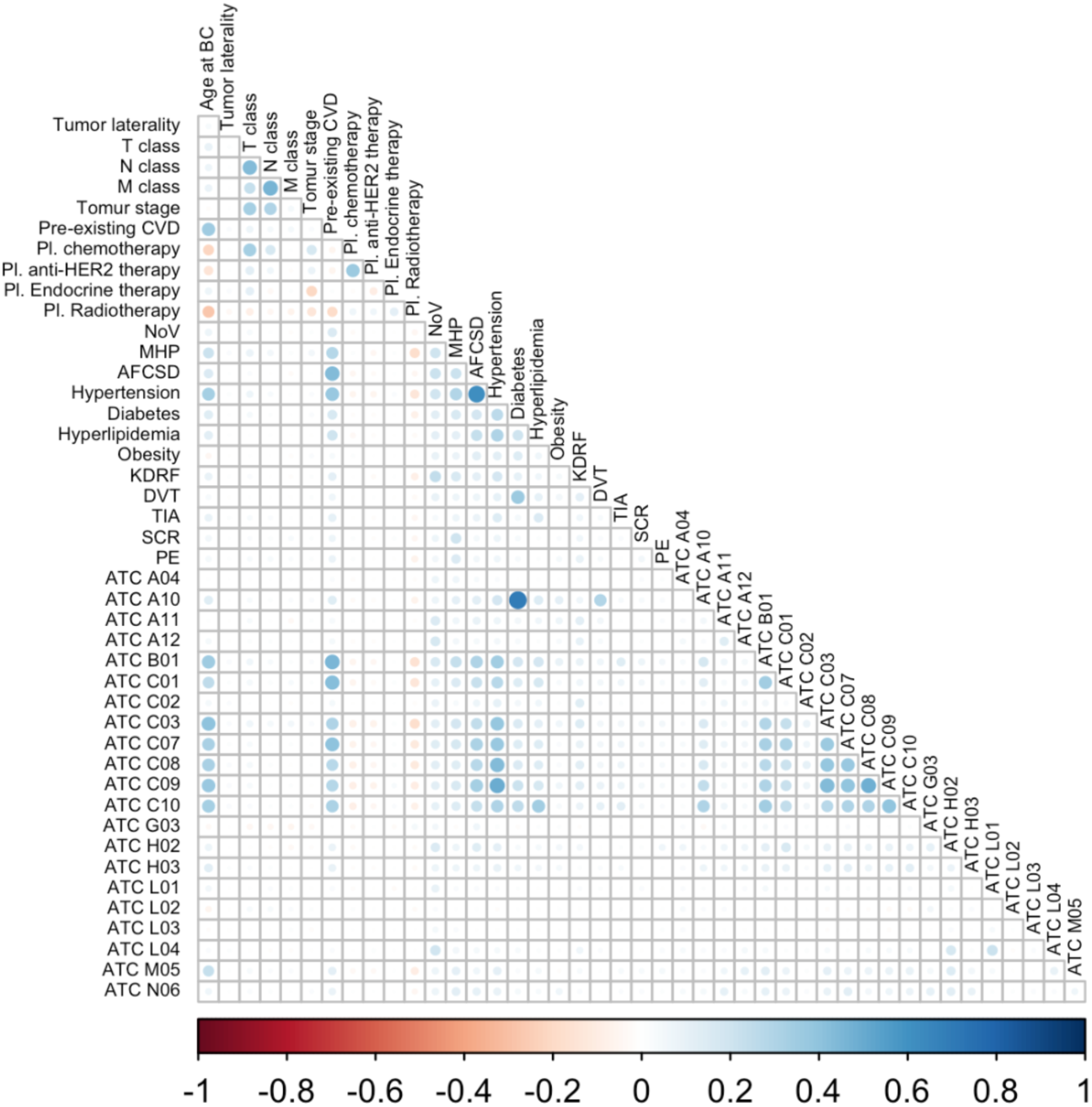
Pairwise Pearson correlation between features. No two features were highly correlated.

### Age contributes to 1-year MACE risk primarily through nonlinear and interaction effects

We passed the top ten important features to a Fine-Gray model to obtain direction and magnitude of associations between covariates and 1-year MACE (Table 2). According to the Fine-Gray model, the age at BC diagnosis, maximum hospitalization period, and number of visits were associated with a modestly elevated risk for MACE; however, they contributed strongly to the predictive performance of our model. Patients with prior CVD records or an unassessable primary tumor stage showed the strongest association with elevated risk among the predictors. The G03 medication family, comprising sex hormones and modulators of the genital system, was associated with a lower risk and was the only predictor showing a protective association.

**Table 2.** The Fine-Gray evaluated hazard ratios (HR) and corresponding p-values for the top ten features of the trained model.

| Predictor | HR | *p*-value |
| --- | --- | --- |
| Age | 1.066 | <0.001 |
| Prior CVD | 1.958 | <0.001 |
| ATC C09 | 1.161 | 0.06 |
| Maximum hospital stay | 1.005 | 0.16 |
| Visit count | 1.005 | 0.06 |
| ATC B01 | 1.075 | 0.42 |
| Chemotherapy | 1.146 | 0.08 |
| ATC G03 | 0.851 | 0.024 |
| Stage II | 1.398 | <0.001 |
| Stage III | 1.252 | 0.24 |
| Stage insitu | 0.619 | 0.18 |
| Stage x | 2.145 | 0.013 |
| ATC C01 | 1.210 | 0.045 |

## Discussion

This study demonstrates that a deep learning-based competing-risk model can accurately predict the short-term risk of MACE in women with BC using pre-treatment data alone. Across three CV folds, the model showed strong and stable discriminative performance for both the primary outcome and the competing event, with consistently high c-index values. These results indicate that the model captures clinically meaningful heterogeneity in individual cardiovascular risk, even in the presence of substantial competing non-cardiovascular mortality, and that its performance is not driven by data partitioning or overfitting.

In contrast to most existing cardiovascular risk prediction tools, our approach is deliberately restricted to information available before initiation of cancer-directed therapy, addressing a critical decision point in routine oncology care. General-population risk scores such as QRISK3, SCORE2, PCP-HF, and CHARGE-AF assume stable long-term risk factor profiles, standardized cardiovascular measurements, and the absence of competing cancer-related mortality, limiting their applicability in newly diagnosed BC patients^4–7^. Cancer-specific cardio-oncology models have advanced the field but typically rely on post-treatment or on-treatment variables, including anthracycline dose, trastuzumab exposure, radiotherapy details, or echocardiographic parameters such as left ventricular ejection fraction or strain ^8,9,10,11,12^.

More recent machine-learning and deep-learning approaches have demonstrated promising discrimination but similarly depend on rich longitudinal EHR data, detailed treatment trajectories, laboratory measurements, imaging, or multimodal inputs that are not routinely available at the initial oncology consultation ^15, 16, 17^. By contrast, our model intentionally excludes echocardiography, ECG-derived features, cardiac biomarkers, and delivered treatment information, and instead leverages demographics, cancer characteristics, baseline comorbidities, medication profiles, and healthcare utilization recorded up to diagnosis. This design allows individualized, time-dependent estimation of MACE risk before systemic therapy is initiated, while explicitly accounting for non-cardiovascular death as a competing risk.

From a methodological perspective, while conventional models such as Fine-Gray regression provide interpretable estimates of marginal associations, they assume proportional subdistribution hazards and linear covariate effects. In contrast, the DeepHit framework of our model allows for nonlinear effects and complex interactions among covariates, which is particularly relevant in heterogeneous clinical populations. The observed discriminative performance suggests that such nonlinear modeling contributes meaningfully to individualized risk prediction. Several predictors, including age at BC diagnosis, maximum hospitalization period, and number of visits, exhibited only modest hazard ratios in the Fine-Gray model yet ranked highly in permutation-based importance analyses within our model. This discrepancy suggests that these variables contribute to risk stratification primarily through nonlinear effects and interactions rather than strong marginal associations. This underscores a key advantage of deep learning-based survival models over traditional regression frameworks.

Apart from the expected strong association of prior CVD with the first-year risk of MACE, the strong association observed for unassessable tumor stage likely reflects underlying disease complexity, delayed diagnosis, or incomplete clinical information, rather than a direct biological effect. In contrast, the G03 medication family, which includes sex hormones and modulators of the genital system, was the only variable associated with a lower predicted cardiovascular risk. This finding should be interpreted cautiously, as it may reflect treatment selection, underlying hormonal milieu, or residual confounding rather than a direct protective pharmacological effect. Biological evidence suggests that estrogen interacts with vascular estrogen receptors to influence endothelial function, vasodilation, and vascular homeostasis, which could have favorable effects on cardiovascular profiles in some contexts ^30,31^.

Observational studies also report a lower incidence of CVD in premenopausal women compared to age-matched men, and an increase in cardiovascular risk following menopause, consistent with the loss of endogenous estrogen exposure ^32^. However, randomized trials of menopausal hormone therapy in older postmenopausal women have yielded neutral or heterogeneous findings with respect to cardiovascular outcomes, indicating that the relationship between hormone exposure and clinical cardiovascular risk is complex and influenced by timing, formulation, and disease state ^33,34^. Thus, although the association of G03 medications with lower predicted cardiovascular risk in our model aligns with some mechanistic and observational data, causal inference cannot be drawn, and further study is needed to disentangle hormonal effects from confounding and treatment indication.

Limitations of this study should be also acknowledged. First, the study is based on observational data, and the identified associations should not be interpreted causally. Second, although CV demonstrated stable performance, external validation in independent cohorts is necessary to assess generalizability. Third, the Fine-Gray analysis was restricted to the most important features identified by our model, and results may differ if alternative feature sets or modeling assumptions are used. Finally, genetic or genomic information was not available in this cohort, which may limit the ability to capture underlying biological heterogeneity relevant to cardiovascular risk.

Taken together, these findings suggest that our model provides a robust framework for individualized cardiovascular risk stratification in oncology patients, effectively accounting for competing risks and complex covariate relationships. By integrating heterogeneous clinical, treatment, and utilization data, the model may support early identification of high-risk individuals who could benefit from targeted surveillance or preventive strategies. The model is therefore positioned to complement, rather than replace, existing cardio-oncology tools by supporting early, risk-based referral decisions in settings with limited cardiology resources.

## Conclusion

In conclusion, this study shows that the short-term risk of MACE in women with BC can be meaningfully assessed using a deep learning-based competing-risk model that relies solely on information available at the time of diagnosis. By accounting for competing non-cardiovascular mortality and allowing for nonlinear relationships among routinely collected clinical and healthcare data, the model offers individualized risk estimates at a point when treatment decisions are being made, and cardiology referral is most relevant. Rather than replacing existing cardio-oncology tools, this approach is intended to complement them by supporting earlier and more targeted referral and surveillance strategies in settings where resources are limited. With further external validation and prospective evaluation, pre-treatment risk assessment of this kind may help improve cardiovascular care and long-term outcomes for women undergoing treatment for breast cancer.

## Data Availability

The data that support the findings of this study are restricted. Access can be requested through the Swedish National Data Service for research purposes, subject to ethical approval.

